# Breast cancer polygenic risk score performance varies by socioeconomic status

**DOI:** 10.64898/2026.06.03.26354819

**Authors:** Henna Domian, Xiaohe Tian, Dylan Ong, Lily Hamilton, Yiwey Shieh, Shaila Musharoff

## Abstract

**Background:** Polygenic risk scores (PRS) for breast cancer are increasingly used for risk stratification to inform screening and prevention. However, for PRSs to be equitable and clinically useful, they need to perform well across diverse populations. While PRS performance is known to be ancestry-dependent, it is not well understood how environmental context, such as that of socioeconomic status (SES), affects PRS transferability. Here, we assess whether SES, measured via self-reported household income, modifies breast cancer PRS performance and, if so, whether socioeconomic context contributes predictive information beyond genetic risk alone.

**Methods:** We used the US-based All of Us biobank to evaluate how SES impacts breast cancer PRS performance. First, we quantified changes in breast cancer PRS performance by modeling a commonly-cited polygenic score for breast cancer previously described by Mavaddat et al. with SES. We then reestimated the genetic effect sizes of the 3,820 variants from Mavaddat et al. in All of Us with and without income as a covariate. Because social determinants of health affect breast cancer detection and outcomes, we stratified analyses by socially defined populations on the basis of self-identified race and ethnicity. We further stratified individuals whose self-identified race is White (“White”) into three SES groups (high, middle, low) based on self-reported income and re-estimated genetic effect sizes to create SES-specific PRSs. We then applied these PRSs to White participants, the largest group in the study, and to Black or African American (“Black”) and Hispanic or Latino (“Hispanic”) participants, groups underrepresented in breast cancer research. Model discrimination between cases and controls was measured by area under the curve (AUC).

**Results:** We analyzed 163,715 women from the All of Us biobank, which included 8,833 breast cancer cases (6,619 White, 1,178 Black, and 1,036 Hispanic), with relative income available for a subset of these cases (5,525 White, 848 Black, and 566 Hispanic). The ancestry-dependent performance of the breast cancer PRS described in Mavaddat et al. was replicated in All of Us. In Black individuals, this PRS (AUC and 95% CI: 0.576 [0.571, 0.582]) produced a similar increase in AUC as relative income (AUC: 0.573 [0.568, 0.577]) when added to an age-only model. Incorporating income with PRS, age, and genetic PCs 1-3 improved AUC by 0.007 in White Americans and 0.018 in Black Americans (both p < 10^-11^), while attenuating the contribution of PRS in the full model. PRS performance also varied among SES categories. Notably, PRSs with variant effect sizes that were recalibrated in low-SES White participants performed best in low-SES White participants (AUC: 0.605 [0.583, 0.628]) and Black Americans (AUC: 0.588 [0.586, 0.591]), both better than performance in high-SES White Americans (AUC: 0.579 [0.577, 0.580]) and middle-SES White Americans (AUC: 0.578 [0.569, 0.586]).

**Conclusion:** Socioeconomic context, measured by income, significantly impacts the transferability of a PRS for breast cancer within and among groups defined by self-identified race and ethnicity. Accounting for SES improves PRS performance, most notably in Black Americans and low-SES White individuals.

## Introduction

Cancer development reflects a complex interplay between genetic risk, environment, and lifestyle. While current breast cancer risk prediction relies on factors including family history and tests for inherited genetic mutations, such as in the BRCA1/BRCA2 genes, they miss heritable risks that whole-genome profiling can reveal (Torkamani et al., 2018). Genome-wide association studies (GWAS) enable the evaluation of the genomes of large samples of people to identify associations between specific genetic variants, including single-nucleotide polymorphisms (SNPs), and specific phenotypes, including disease. Polygenic risk scores (PRS) aggregate the effect of trait-associated genetic variants identified by GWAS to predict complex trait values. PRS can improve disease risk assessment, including for breast cancer (Maas et al., 2016; Schumacher et al., 2018). Recent prospective studies have highlighted the utility of PRS in tailoring breast cancer screening and prevention on a population level (Esserman et al., 2026). In the WISDOM Study, integration of breast cancer PRS changed screening recommendations for a subset of participants, increasing screening recommendations for some women and decreasing them for others (Fergus et al., 2025). Modeling studies have also suggested that breast cancer screening strategies incorporating PRS and family history can avert more breast cancer deaths than family-history-based approaches alone (van den Broek et al., 2021). PRS can also stratify risk among BRCA1 and BRCA2 pathogenic variant carriers, suggesting potential utility for individualized risk management in genetically high-risk populations (Kuchenbaecker et al., 2017).

Genetic ancestry is a central consideration in PRS research because European-derived scores often transfer poorly across populations. European-derived PRS for breast cancer perform better in those of European ancestry (Area under the Curve, AUC of 0.63) compared to those of African ancestry (AUC 0.53) (Mavaddat et al., 2019; Liu et al., 2021). Previous studies have attributed differences in PRS transferability between populations to differences in population allele-frequency variation, linkage disequilibrium structure, and demographic history (Pritchard and Przeworski, 2001; Martin et al., 2017; Kachuri et al., 2024). Because most PRS have been developed and validated in European-ancestry populations, their clinical use may widen existing health disparities unless performance is improved in diverse populations (Duncan et al., 2019; Martin et al., 2019; Wang et al., 2022). Recent work has addressed this gap directly. Li et al. developed improved PRS for overall and subtype-specific breast cancer in African ancestry women, including estrogen receptor-positive, and triple-negative breast cancer (Li et al. 2026). Shieh et al. evaluated a breast cancer PRS for US Latin American women, showing that risk models can stratify breast cancer risk in Hispanic populations with diverse ancestry (Shieh et al., 2020). These efforts are important because ancestry-related differences in PRS performance can overlap with disparities experienced by socially defined racial and ethnic groups. This is further confounded with socioeconomic status, reflecting environmental exposures and how early breast cancer risk is detected. Triple-negative breast cancer is more common among Black women than White women and is associated with poorer disease outcomes (Dietze et al., 2015; Prakash et al., 2020). Hispanic and Latina women experience disparities with breast cancer screening, resulting in higher overall mortality, despite having a lower incidence for breast cancer (Serrano-Gómez et al., 2020). Together, these findings underscore the need to evaluate whether PRS models perform equitably across populations shaped by both genetic ancestry and social determinants of health.

Researchers have also made a considerable effort to increase the number of non-White individuals being sampled to address this. The All of Us Research Program is a United States NIH initiative that collected and made accessible biobank-scale genetic, phenotypic, and health data, and prioritized diversity by including populations historically underrepresented in genomic research. The latest Controlled Data Repository version 8 (CDRv8) release includes short-read whole-genome sequencing (WGS), demographic, clinical, and environmental data for 414,830 participants, 190,833 of whose self-reported race is something other than “White” (All of Us, 2025).

Neighborhood socioeconomic status (nSES) has been implicated in breast cancer risk, particularly for aggressive subtypes. Prior studies have found that lower nSES is positively associated with the incidence of triple-negative and other aggressive breast cancer subtypes, with these patterns especially relevant among Black women (Qin et al., 2021; Aoki et al., 2021). Because nSES captures a composite of environmental, structural, and healthcare-related exposures, it may contribute to differences in breast cancer risk that are not explained by inherited genetic variation alone. Polygenic score performance is also context dependent. Mostafavi et al. showed that PGS prediction accuracy can vary across social and demographic strata, including socioeconomic strata defined by the Townsend deprivation index (Mostafavi et al., 2020). Recent work has emphasized that PRS implementation should account for social determinants of health because genetic and environmental risk vary across populations (Cromer et al. 2026). For cancer specifically, studies have begun to test how genetic risk varies across socioeconomic context, including prostate cancer risk within the All of Us biobank (Cheng et al. 2026). However, relatively little work has evaluated whether socioeconomic context can be used to improve breast cancer PRS performance. This motivates testing whether SES-adjusted or SES-stratified recalibration improves breast cancer PRS transferability across ancestry groups.

In this work, using the All of Us Research Program data, we assess whether socioeconomic context as measured by self-reported income contributes to PRS transferability and the predictive power of PRS for breast cancer beyond genetic risk alone. We specifically test whether the socioeconomic context of a training group affects model performance across groups defined by socioeconomic strata and self-identified race and ethnicity, beyond age, PRS, and genetic ancestry.

## Methods

### Overall Design

We conducted complementary analyses in All of Us to assess socioeconomic effects on breast cancer polygenic score performance. First, we conducted an additive covariate analysis using fixed, previously estimated PRS weights from Mavaddat et al. to quantify the incremental predictive value of socioeconomic and demographic variables in age-matched case-control datasets (Mavaddat et al., 2019). Second, we created a recalibrated PRS in which variant effect sizes were re-estimated within White participants. PRS were recalibrated in non-stratified and SES-stratified White individuals where we utilized household income relative to the federal poverty line as a measure of SES. In both analyses, we assessed the impact of model modifications on discrimination, as measured by area under the curve (AUC).

### Study Population

We analyzed participants from the All of Us biobank (v8) whose self-identified gender is “Woman”, whose sex assigned at birth is “Female”, and who have available whole-genome sequencing and electronic health record (EHR) data. We stratified our sample first by self-identified race and ethnicity, and later by self-reported income relative to the federal poverty line. This allowed us to evaluate PRS performance across socially defined groups who may experience different structural exposures, including environmental exposure and healthcare access. Genetic ancestry was accounted for with genetic principal components (PCs) adjustments, rather than using genetic ancestry to define groups. We limited our analysis to those whose self-identified race and ethnicity as reported on the the All of Us Basics survey is either “White” (here, “White”), “Black or African American” (here, “Black”), or “Hispanic or Latino” (here, “Hispanic”). These are the three largest non-overlapping identities in the All of Us Cohort Builder’s self-reported category variable. This yielded 163,715 individuals with genetic data (99,325 White; 31,582 Black; 32,808 Hispanic), of which 8,833 are cases with a prior diagnosis of breast cancer (6,619 White; 1,178 Black; 1,036 Hispanic). Breast cancer case status was defined as malignant neoplasm of the breast using SNOMED concept ID 4112853 and encoded as a binary variable. Analyses involving socioeconomic variables were restricted to participants who responded to the household income and family size questions in the All of Us Basics survey. This left 128,132 individuals with genetic, EHR, and income data (85,556 White; 23,358 Black; 19,218 Hispanic), of which 6,939 are cases with a prior diagnosis of breast cancer (5,525 White, 848 Black, and 566 Hispanic) [Fig. 1].

**Figure 1.**
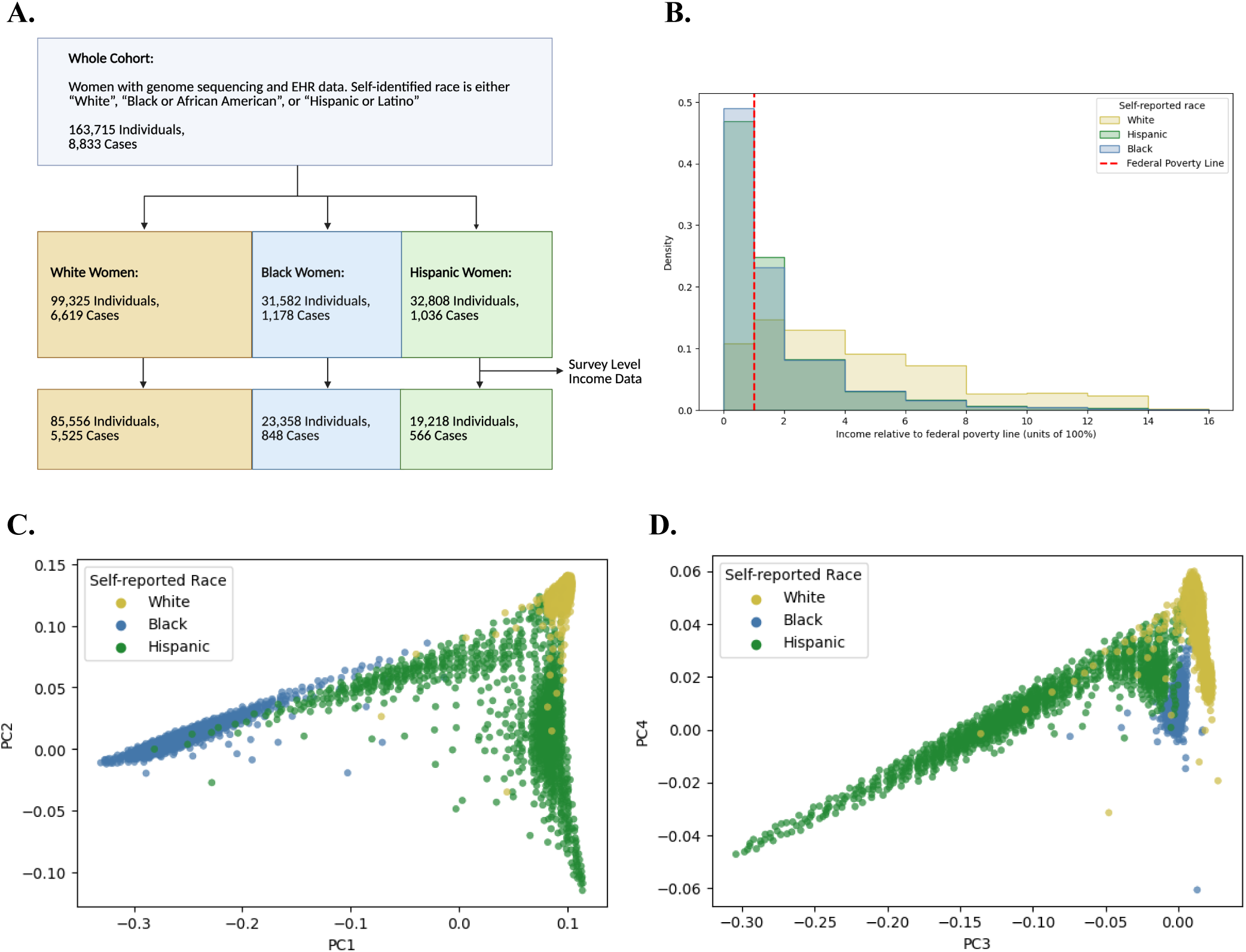
Income, ancestry, and self-reported racial and ethnic identity distributions of groups in *All of Us.* (B) Cohort selection and sample sizes for women with genome sequencing and EHR data, stratified by self-identified race and ethnicity and then restricted to participants with survey-level household income and family size data. (A) Income relative to the federal poverty line in units of 100 percent for 85,556 White American women is significantly higher than that of the 23,358 Black and 19,218 Hispanic Americans. (C) Genetic PCs 1 and 2 colored by self-reported race. (D) Genetic PCs 3 and 4 colored by self-reported race. Each point in (B) and (C) represents a centroid of 30 individuals.

### Socioeconomic measures

Household income relative to the federal poverty line (“relative income”) was calculated based on the year in which each participant completed The All of Us Basics survey between 2017 and 2025. For each survey year, we used the federal poverty guideline base value and per-person increment for the 48 contiguous states and the District of Columbia, as reported by the Department of Health and Human Services. These values ranged from $12,060 + $4,180 × [household size − 1] in 2017 to $15,650 + $5,500 × [household size − 1] in 2025 (Department of Health and Human Services, 2025). Household income was self-reported in discrete intervals on the survey, and we used the midpoints of these intervals as income. We then expressed income as a percentage of the federal poverty line, in units of 100%, to calculate relative income. We obtained three-digit ZIP code neighborhood deprivation from All of Us. The deprivation index was based on a principal components analysis of six different socioeconomic variables from the 2023 American Community Survey (ACS) (Brokamp et al., 2019; Brokamp, 2023).

### Polygenic Scoring and Genetic Data

We obtained 3,820 breast cancer risk-associated variants (3,220 SNP and 600 insertion/deletion) identified by Mavaddat et al. from the PGS catalog (Mavaddat et al., 2019; Lambert et al., 2021). We then performed a LiftOver operation using the UCSC genome browser from GRCh37 to GRCh38 (UCSC Genome Browser, 2021). After restricting to All of Us ACAF threshold variants (defined as having population-specific allele frequency (AF) > 1% or allele count (AC) > 100), 3,683 variants (3,148 SNP and 535 insertion/deletion) were mapped to the All of Us dataset. We then calculated a breast cancer PRS with PLINK 1.9 as PRS = β_1_\**x*_1_ +β_2_\**x*_2_+ … +β_*n*_\**x*_*n*_, where β_i_ is the log of the odds ratio (OR) for the breast cancer-associated variant x_i_ for each of *n* variants (Chang et al., 2015). We normalized the PRS to the mean and standard deviation in women without a breast cancer diagnosis (“controls”) within self-identified racial groups (Black, White, or Hispanic). We obtained genetic principal components (PCs) as calculated by All of Us (The All of Us Research Program Genomics Investigators, 2024) and clustered them with a minimum cluster size of 30 individuals for visualization purposes.

### Additive Covariate Analysis

To assess the effect of covariates on breast cancer risk prediction, we fit sequential additive linear models. Within each racial group, we selected 5 age-matched controls for each case by sampling without replacement from age-matched controls. Controls were selected from 5-year age bins spanning 20-110 years (which is nearly identical to the case age range of 23-106 years) [Supplementary Fig. 1]. Age matching was repeated across 10 independent repetitions. Previously estimated PRS weights were used without modification. Logistic regression models were evaluated using 5-fold cross-validation repeated across these 10 repetitions. We assessed the performance of the following sequential additive models in predicting breast cancer: Age; Age + Relative Income; Age + PRS; Age + PRS + PCs 1-3; Age + PRS + PCs 1-3 + Relative Income. Discrimination was quantified by AUC. An AUC of 0.5 indicates no discriminative ability, while an AUC of 1.0 represents perfect discrimination.

### 5-Fold Cross-Validation for PRS variants

We recalibrated the PRS originally derived by Mavaddat et al. by re-estimating the effect size of each variant in All of Us in samples specified below, using a 5-fold cross-validation framework with repeated down-sampling. Cases were partitioned into five folds, with four folds (80%) used for training and one fold (20%) reserved for validation. For each fold, controls were sampled without replacement from a large control pool (described below) to achieve a 1:5 case-to-control ratio in the training set. Each fold was repeated 10 times with a newly sampled set of controls to account for variability introduced by this down-sampling. Cases were fixed across repeats. Within each fold repetition, PLINK logistic regression was used to re-estimate variant effect sizes, and variants reaching p < 0.05 were used to construct a recalibrated PRS. Model performance was assessed in the held-out validation fold using a 1:5 case-to-control ratio. For each validation fold, the 10 AUCs calculated from reselecting controls were first averaged to generate one fold-level AUC. Final performance was reported as the mean AUC and 95% CI across the five fold-level AUCs. A schematic of the PRS recalibration and evaluation workflow for a single fold-resample iteration is shown in Figure 4.

### PRS recalibration

To determine whether training with income improves PRS performance, we used the 5-fold cross-validation framework (described above) with 128,132 White, Black, and Hispanic individuals with genetic and income data. We recalibrated PRSs in White All of Us participants by re-estimating β-effect weights for each PRS variant using logistic regression in PLINK. Two training sets were evaluated: (i) 1,500 cases and 15,000 controls and (ii) 5,525 cases and 80,031 controls. The smaller size matched the case counts used in the SES-stratified analyses below, and the larger sample size selected all White American cases with survey level income data. We evaluated three model specifications: PRS only, PRS trained with relative income included during variant effect size estimation but excluded at evaluation, and PRS with relative income included both during training and as a covariate in the model. Transferability of the PRS between different groups was evaluated by applying PRSs trained in White participants separately to the Hispanic (566 cases, 18,652 controls) and Black (848 cases, 22,510 controls) cohorts. We summarized predictive performance as the mean AUC across folds and resamples [Fig. 5].

### SES-stratified PRS recalibration

We then repeated the 5-fold cross-validation framework (described above) in White participants who were stratified by relative income. We did this to assess whether income-specific effects differ across low, middle, and high SES training cohorts, defined as having relative income that is <300%, 300–600%, and >600% of the federal poverty line, respectively. Cases were down-sampled to 1,500 cases per stratum to ensure comparability across groups (cases: 2,140 high, 1,868 middle, and 1,517 low income), with a 15,000 control resampling pool per stratum. Relative income was included in training and as a covariate in the final prediction model. In each fold, controls were resampled 10 times, resulting in training folds having 1,200 cases and 6,000 controls. We then evaluated PRSs in held-out validation folds (300 cases and 1,500 controls) with relative income included as a covariate. We tested SES-specific PRSs (low, middle, or high) in independent SES-matched (stratified) White cohorts. We also tested them in SES-unstratified Black (848 cases, 22,510 controls) and Hispanic (566 cases, 18,652 controls) cohorts [Fig. 6].

## Results

### Characteristics of the study population

Our analysis included 163,715 individuals, of whom 8,833 were previously diagnosed with breast cancer (6,619 White, 1,178 Black, and 1,036 Hispanic individuals). After restricting to those who answered survey questions regarding family size and household income, 128,132 individuals remained, 6,939 of whom were previously diagnosed with breast cancer (5,525 White, 848 Black, and 566 Hispanic cases) [Fig. 1]. Figure 1 also shows the distribution of relative income and clustered genetic PCA results for the analysis set of individuals stratified by self-reported race.

### Performance of the published breast cancer PRS in All of Us

PRS distributions for each group are shown in Figure 2. This score applied to the All of Us dataset replicated previously reported performance, as accuracy as measured by area under the receiver operating characteristic curve (AUC) of 0.617 in White Americans, 0.577 in Black Americans, and 0.573 in Hispanic Americans, demonstrating reduced predictive performance in non-White groups. An AUC of 0.5 indicates no discriminative ability, while an AUC of 1.0 represents perfect discrimination.

**Figure 2.**
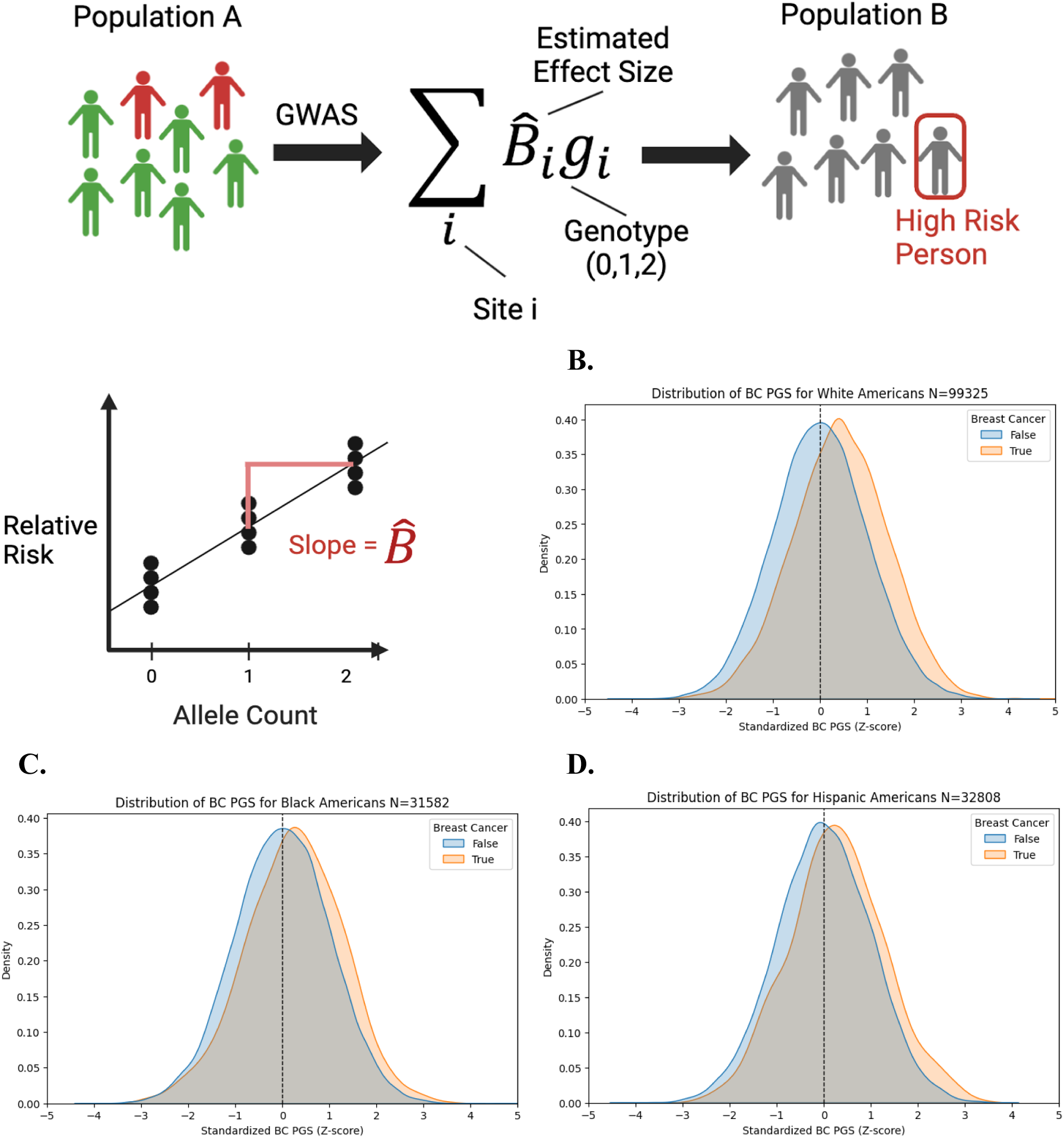
Translation of Breast Cancer PRS from Mavaddat et al. (2019) to All of Us biobank, normalized on controls for each population. (A) GWAS and PRS construction schematic. The distribution of this score is shown in (B) White Americans (6,619 cases, 99,325 controls), (C) Black Americans (1,178 cases, 31,582 controls), and (D) Hispanic Americans (1,036 cases, 32,808 controls).

### Additive covariate models incorporating socioeconomic status

Figure 3 shows AUC distributions for additive covariate models incorporating age, relative income, PRS, and genetic PCs 1–3, stratified by self-identified race and ethnicity. Table 1 shows odds ratios (ORs) with confidence intervals, effect sizes, and p-values per racial group for each variable modeled in Figure 3. Supplementary Table 1 provides fold- and repeat-level AUCs, training and validation sample sizes, and mean ages for each additive covariate model.

**Figure 3.**
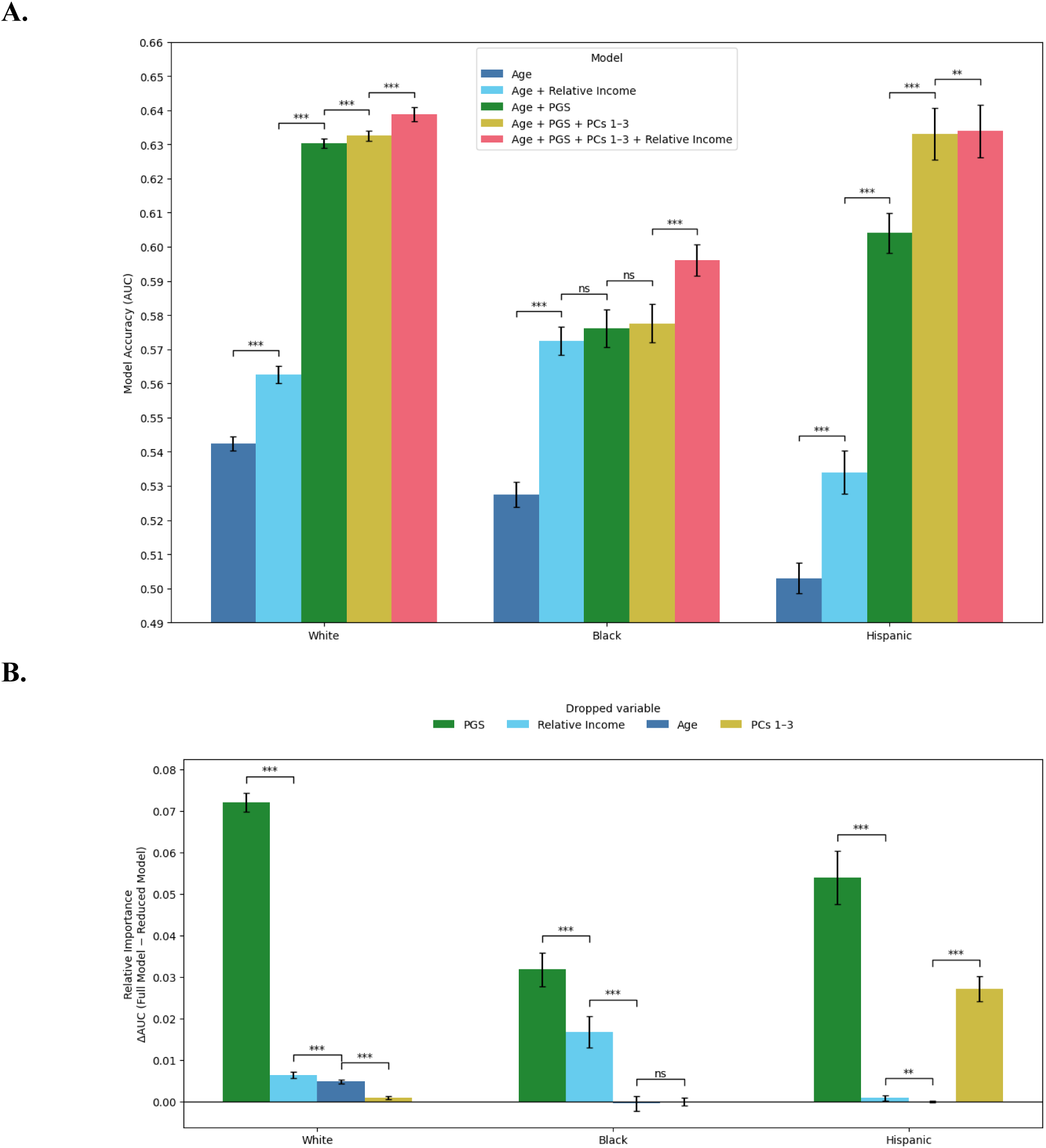
Model performance (AUC) for predicting breast cancer status using additive models across groups. (A) Bars represent 5-fold cross-validated AUCs with a 1:5 case to control ratio across 10 independent repetitions for White (left), Black (middle), and Hispanic Americans (right), sampled from an age-matched dataset [Supplementary Fig. 1]. Models are additive: Age; Age + Relative Income; Age + PRS; Age + PRS + Genetic PCs 1-3; and Age + PRS + PCs 1-3 + Relative Income. The PRS uses previously published weights without recalibration. (B) Change in AUC as a result of removing each covariate from the complete model. Error bars denote 95% CIs around the mean AUC, and 1-3 significance stars representing cutoffs of p < 0.05, p < 0.01, and p < 0.001 via paired t-tests.

**Table 1.** Odds ratios and a 95% confidence interval, p-values, and effect sizes for all covariates included in the full breast cancer risk model, stratified by self-identified race and ethnicity.

| Covariate | Odds Ratio | P-Value | Effect Size |
| --- | --- | --- | --- |
| **White Americans** |  |  |  |
| Breast Cancer PRS | 1.55 [1.51, 1.59] | $1.38 \times 10^{-197}$ | 0.43 |
| Relative Income | 1.17 [1.14, 1.20] | $4.62 \times 10^{-30}$ | 0.16 |
| Age | 1.14 [1.11, 1.18] | $2.10 \times 10^{-19}$ | 0.13 |
| PC1 | 1.04 [1.00, 1.08] | $7.77 \times 10^{-02}$ | 0.04 |
| PC2 | 0.94 [0.92, 0.98] | $4.20 \times 10^{-04}$ | -0.06 |
| PC3 | 1.06 [1.03, 1.10] | $4.84 \times 10^{-04}$ | 0.06 |
| **Black Americans** |  |  |  |
| Breast Cancer PRS | 1.34 [1.24, 1.44] | $4.33 \times 10^{-15}$ | 0.29 |
| Relative Income | 1.18 [1.11, 1.26] | $1.84 \times 10^{-07}$ | 0.17 |
| Age | 1.07 [1.00, 1.15] | $6.52 \times 10^{-02}$ | 0.07 |
| PC1 | 0.90 [0.71, 1.14] | $3.65 \times 10^{-01}$ | -0.11 |
| PC2 | 1.15 [0.91, 1.47] | $2.46 \times 10^{-01}$ | 0.14 |
| PC3 | 1.02 [0.93, 1.11] | $6.86 \times 10^{-01}$ | 0.02 |
| **Hispanic Americans** |  |  |  |
| Breast Cancer PRS | 1.50 [1.37, 1.64] | $4.96 \times 10^{-19}$ | 0.40 |
| Relative Income | 1.07 [0.98, 1.16] | $1.36 \times 10^{-01}$ | 0.06 |
| Age | 0.98 [0.90, 1.07] | $6.86 \times 10^{-01}$ | -0.02 |
| PC1 | 0.84 [0.61, 1.14] | $2.57 \times 10^{-01}$ | -0.18 |
| PC2 | 1.53 [0.86, 2.74] | $1.51 \times 10^{-01}$ | 0.43 |
| PC3 | 0.87 [0.43, 1.76] | $6.92 \times 10^{-01}$ | -0.14 |

Age alone had modest predictive value, with mean AUCs of 0.542 [0.540, 0.544] in White participants, 0.528 [0.524, 0.531] in Black participants, and 0.503 [0.499, 0.507] in Hispanic participants. In the full model, age was significantly associated with breast cancer only in White participants, with an OR of 1.14 [1.11, 1.18], consistent with the larger age discrepancy between cases and controls in this cohort [Supplementary Fig. 1, Table 1, Supplementary Table 1]. Adding relative income to age improved discrimination in all groups, with the largest gain observed in Black participants. Mean AUC increased to 0.563 [0.560, 0.565] in White participants, 0.573 [0.568, 0.577] in Black participants, and 0.534 [0.528, 0.540] in Hispanic participants. In the full model, relative income was associated with breast cancer in White and Black participants, with ORs of 1.17 [1.14, 1.20] and 1.18 [1.11, 1.26], respectively [Table 1]. Incorporation of the published PRS to a base model with age produced the largest gain in discrimination. Mean AUC increased to 0.630 [0.629, 0.632] in White participants, 0.576 [0.571, 0.582] in Black participants, and 0.604 [0.598, 0.610] in Hispanic participants. The PRS was also the strongest predictor in the full model across all groups, with ORs of 1.55 [1.51, 1.59] in White participants, 1.34 [1.24, 1.44] in Black participants, and 1.50 [1.37, 1.64] in Hispanic participants [Table 1]. In Black participants, however, the AUC for age plus relative income was not significantly different from the AUC for age plus PRS by paired t-test.

The full additive model (Age + PRS + PCs 1-3 + Relative Income) achieved the highest AUC in each group. Mean AUC was 0.639 [0.637, 0.641] in White participants, 0.596 [0.592, 0.601] in Black participants, and 0.634 [0.626, 0.642] in Hispanic participants [Fig. 3]. Inclusion of deprivation index did not improve model performance [Supplementary Fig. 2]. The deprivation index in All of Us is assigned at the three-digit ZIP code resolution, which encompasses geographically and socioeconomically heterogeneous populations. The distribution of three-digit zip code deprivation index in our cohort and the relationship between this deprivation index and SES-group are described in Supplementary Figure 3.

### Transferability of All of Us recalibration

Using the PRS recalibration and cross-ancestry evaluation framework [Fig. 4], we evaluated transferability of recalibrated PRSs trained in White participants. Performance varied across populations, with discrimination in Hispanic and Black cohorts differing from that observed in White validation sets. Including relative income as a covariate improved AUC and reduced AUC variability across folds. Inclusion of relative income as a covariate increased AUC in White and Black American participants [Fig. 5]. Paired improvements in AUC after inclusion of relative income for both the 1,500-case and 5,525-case White training sets are shown in Table 2. Fold- and repetition-level AUCs, number of retained variants, case and control counts are tabulated in Supplementary Table 2.

**Figure 4.**
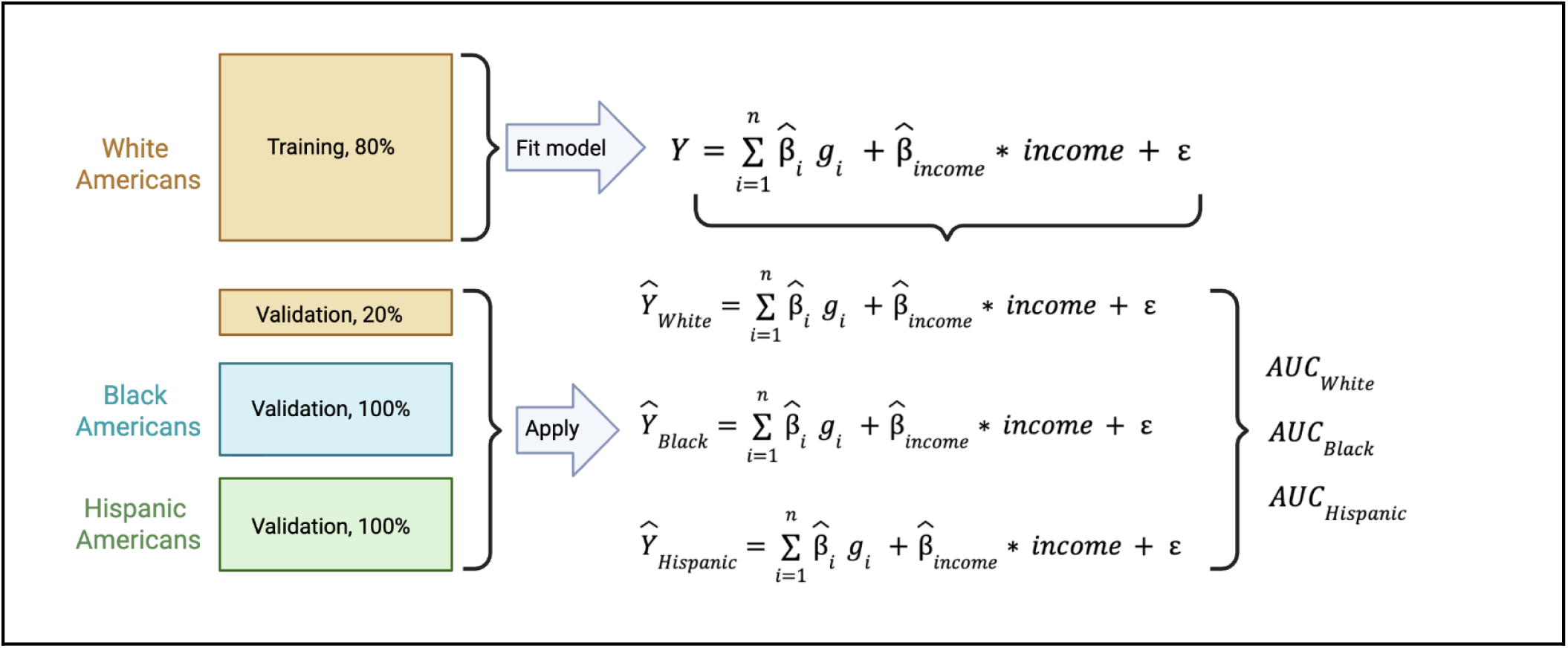
Schematic for k-fold PRS recalibration and cross-ancestry evaluation. White participants were split into training (80%) and validation (20%) sets. Within the training set, variant effect sizes (β^*_i_*) and relative income effect size (β^*_income_*) were estimated using logistic regression. The fitted model was then applied without re-estimation to the held-out White validation set and to the full Black and Hispanic cohorts to generate predicted risk (Ŷ). Predictive performance was quantified as AUC separately in each group. This schematic shows one iteration of the full framework, which is repeated across 5 cross-validation folds, with 10 independent control re-sampling iterations per fold (50 model fits). Results are summarized as the mean AUC across all fold–resample combinations.

**Figure 5.**
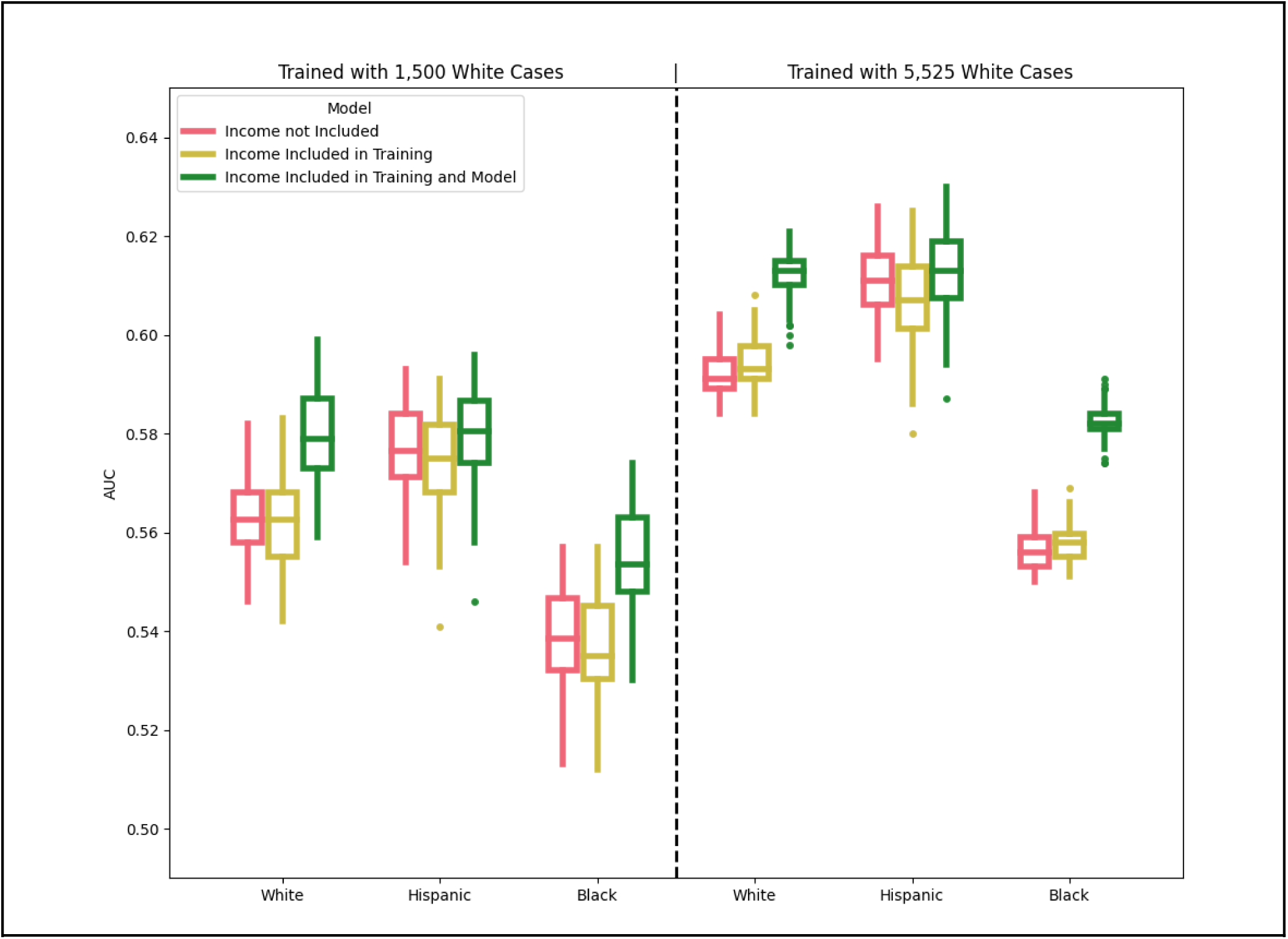
PRS for breast cancer with weights recalibrated in White Americans shows varying predictive accuracy across racial groups. Models include: PRS only (red), PRS with relative income included during variant effect estimation (yellow), and PRS with relative income included both during variant effect estimation and as a covariate in the final model (green). Inclusion of household income relative to the federal poverty line (Income) as a covariate increases model discrimination in White and African American validation cohorts. Five-fold cross-validation was performed with an 80:20 training-validation split, bootstrapped 10 times (N = 50 evaluations per group). PRSs were trained in White Americans using either 1,500 cases and 15,000 controls (left) or 5,525 cases and 80,031 controls (right), and evaluated in White, Hispanic (566 cases, 18,652 controls), and Black (848 cases, 22,510 controls) American validation cohorts. Inclusion of relative income significantly improved AUC in all groups. Larger training sample size on the right resulted in higher overall AUC and tighter distributions across bootstrap replicates.

**Table 2.**
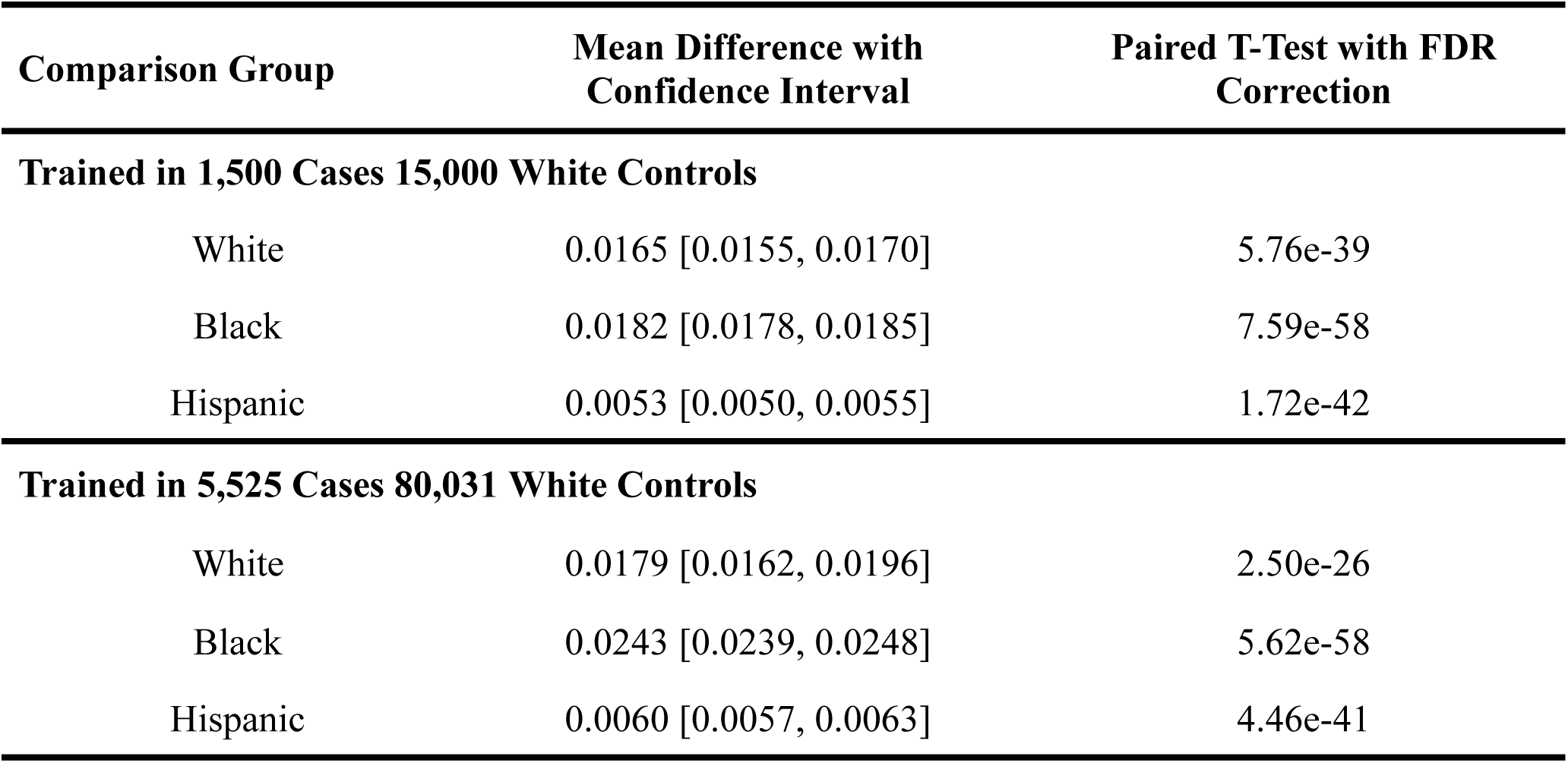
Paired Improvement in Breast Cancer Risk Detection (ΔAUC) After Inclusion of Household Income Relative to the Federal Poverty Line as a Covariate in All of Us PRS Models (1,500 vs 5,525 Case Sets), N=50 (10 bootstraps x 5 folds)

Supplementary Figure 4 extends Figure 5 by including age, relative income, and genetic PCs 1–3 in variant effect-size estimation recalibration. The persistence of AUC changes when relative income is added to the final prediction model indicates that income’s effect on model performance is not explained by genetic ancestry adjustment alone.

### Transferability of SES-stratified recalibration

Figure 6 shows the distribution of AUCs across 50 evaluations from five validation folds and 10 repeated control down-sampling iterations per fold. For summary statistics, the 10 AUCs within each validation fold were first averaged to generate one fold-level AUC. Final performance was reported as the mean AUC and 95% CI across the five fold-level AUCs, reducing inflation of precision from treating repeated down-sampling iterations as independent folds. Fold- and repetition-level AUCs, number of retained variants, income effect estimates, and income p-values are tabulated in Supplementary Table 3.

**Figure 6.**
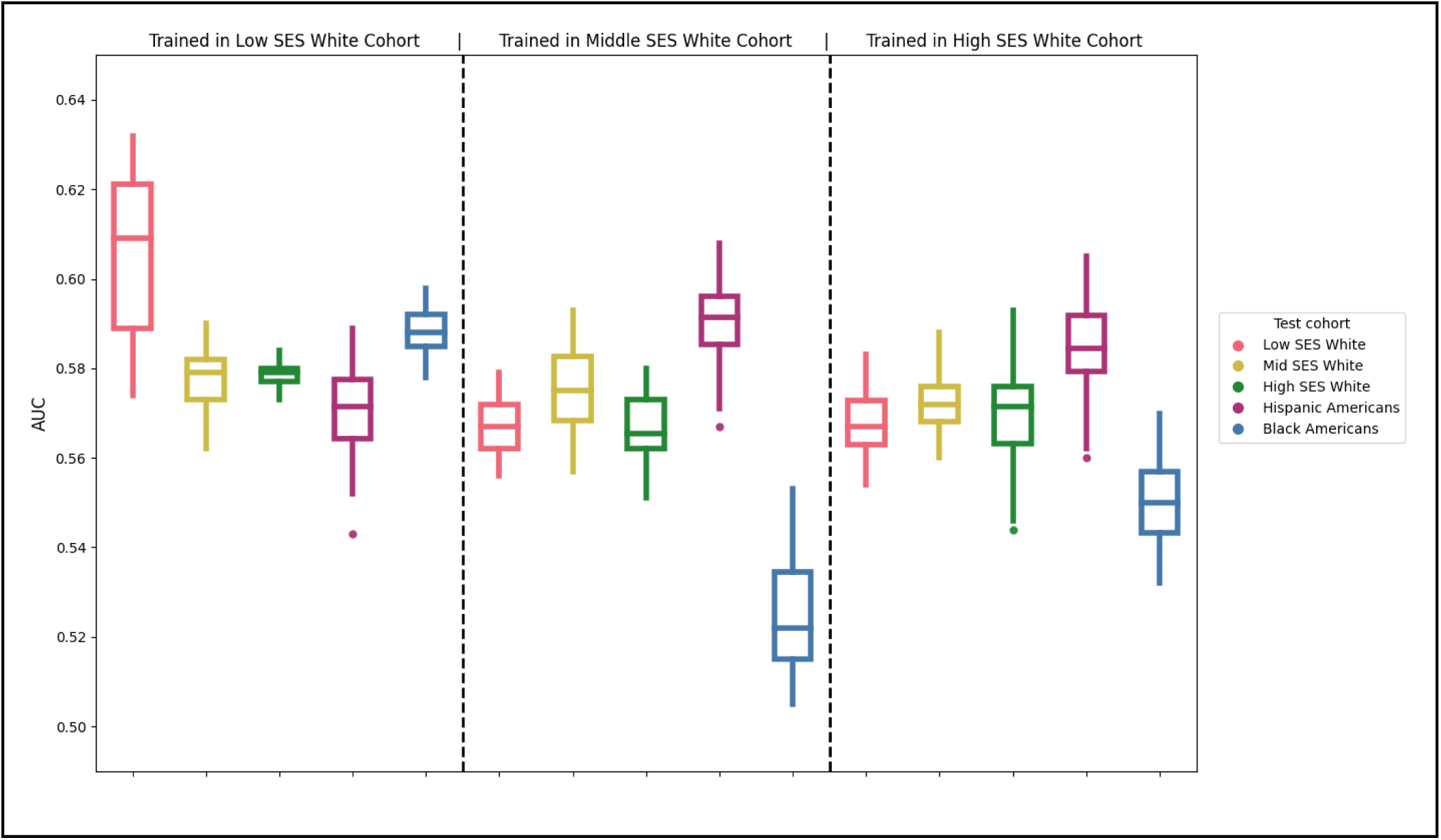
The transferability of White-derived PRS varies by socioeconomic status. White individuals were stratified by household income relative to the federal poverty line into low (<300%; left), middle (300%-600%; middle), and high (>600%; right) SES groups. For each stratum, PRSs were trained using 5-fold cross-validation (1:5 case to control ratio) with relative income included as a covariate during model fitting to estimate variant effect sizes. The resulting SES-stratified PRSs were evaluated in held-out White cohorts stratified by SES and in unstratified Hispanic (566 cases, 18,652 controls) and Black (848 cases, 22,510 controls) cohorts, with relative income included as a covariate in the prediction model. The white SES-stratified training groups consisted of 1,500 cases and 15,000 controls. Significant differences between boxplots are reported in Table S2.

In White Americans, PRSs showed SES-specific transferability. The low-SES-trained PRS had the highest mean AUC in low-SES White participants, at 0.605 [0.583, 0.628], compared with 0.578 [0.569, 0.586] in middle-SES White participants and 0.579 [0.577, 0.580] in high-SES White participants. The middle-SES-trained PRS had the highest mean AUC in middle-SES White participants, at 0.576 [0.566, 0.585], although CIs overlapped with performance in low- and high-SES White participants. The high-SES-trained PRS showed little SES-specific separation, with mean AUCs of 0.568 [0.561, 0.574], 0.572 [0.567, 0.578], and 0.570 [0.559, 0.582] in low-, middle-, and high-SES White participants, respectively.

Transferability to non-White cohorts varied by the SES group used for recalibration. In Black Americans, the low-SES-trained PRS had the highest mean AUC, at 0.588 [0.586, 0.591]. This was numerically higher than the performance of the same PRS in middle-SES White participants, at 0.578 [0.569, 0.586], and high-SES White participants, at 0.579 [0.577, 0.580]. By contrast, PRSs trained in middle- and high-SES White participants showed lower discrimination in Black Americans. Hispanic Americans showed a different pattern, with the highest mean AUC observed for the middle-SES-trained PRS, at 0.591 [0.585, 0.597], followed by the high-SES-trained PRS at 0.585 [0.580, 0.590] and the low-SES-trained PRS at 0.571 [0.563, 0.579]. Including age and genetic PCs 1–3 during variant effect size estimation had minimal impact on the observed patterns [Supplementary Fig. 5]. Supplementary Figure 6 is analogous to Figure 6 but does not include relative income in variant effect size estimation or the final model. Supplementary Figure 7 shows PRSs retrained using the same k-fold framework as Figure 6, but stratified by three-digit ZIP code-level deprivation index rather than survey-derived relative income. Under this coarser ZIP code-based stratification, the SES-related performance trends observed with relative income stratification disappeared: PRSs trained in the low-SES stratum no longer showed differential performance in low-SES White cohorts or in Black cohorts, likely due to the reduced resolution of the deprivation index relative to self-reported income, despite the larger sample size.

## Discussion

In this study, we evaluated the relationship between socioeconomic status and breast cancer polygenic score performance within and across self-reported racial groups using the All of Us biobank. Additive covariate modeling demonstrated that self-reported income relative to the federal poverty line improved discrimination beyond age, genetic ancestry principal components, and a published polygenic score, particularly in Black Americans. The consistent increase in AUC with the inclusion of relative income in our PRS retraining indicates that socioeconomic context contributes discriminatory information beyond genetic risk alone. In addition, our SES-stratified recalibration of a breast cancer PRS showed that transferability depends on the socioeconomic context of the training population. PRSs trained in low-SES White cohorts transferred better to both low-SES White and to Black participants. Both of these groups represent lower-income strata within All of Us (Fig. 1A). This indicates that PRS performance depends on the socioeconomic composition of the training data. Furthermore, socioeconomic context contributes risk-relevant information beyond PRS and genetic PCs.

Our results indicate that socioeconomic context is an additional contributor to variability in PRS performance. Income is correlated with lifestyle, environmental exposures, healthcare access, and other risk-relevant factors that are unevenly distributed across racial groups in the United States. Reduced PRS transferability across genetic ancestry groups is well established, with prior work implicating allele-frequency differences, linkage disequilibrium, demographic history, and underrepresentation of non-European populations in genomic studies (Duncan et al., 2019; Martin et al., 2019; Ding et al., 2023; Kachuri et al., 2024). More broadly, our findings align with evidence that stratifying populations by social or demographic factors can reveal variability in PRS accuracy. Recent work has shown that PRS performance can vary across biobanks and across factors such as age, sex, and income (Mostafavi et al., 2020; Hou et al., 2024). Our findings extend this context-dependent framework to breast cancer risk prediction by showing that relative income contributes predictive information beyond PRS and ancestry PCs. We further show that SES is not only a source of performance variability, but can be leveraged during recalibration, as PRSs trained in low-SES White participants transferred better to both low-SES White and Black American participants.

As the National Academies of Sciences, Engineering, and Medicine recommends, we treat race and ethnicity as social descriptors instead of proxies for genetic ancestry (National Academies of Sciences, Engineering, and Medicine, 2023). This is particularly important here as race and ethnicity vary with socioeconomic status, reflecting environmental exposures and how early breast cancer is detected. Stratified analyses were therefore used to evaluate whether SES-informed PRS models performed equitably across socially defined populations, not to infer biological differences between racial or ethnic groups.

There are several limitations of this study that should be considered. Income was self-reported in brackets and converted to midpoints, introducing potential measurement error in household income relative to the federal poverty line (relative income). Breast cancer case status derived from electronic health records may reflect differences in screening, diagnosis, and healthcare access, which are plausibly correlated with socioeconomic status and could inflate the apparent predictive contribution of socioeconomic variables. Moreover, self-reported income on the study questionnaire was cross-sectional and may not reflect income in the years preceding, or at the time of, breast cancer diagnosis. Restricting analyses to participants who completed the survey questions about income and family size, as we did here, may also introduce selection bias.

The SES-stratified recalibration approach has additional methodological limitations. Re-estimating variant effect sizes involves down-sampling to a specific case-control ratio and filtering variants based on their statistical significance, which may increase sampling variability and produce cohort-specific feature selection. Furthermore, due to the smaller sample sizes for Black and Hispanic Americans in the All of Us v8 dataset, we were unable to apply the SES-stratified recalibration approach that we used in White Americans. All of Us is not a random sample of the United States, and participants contributing data may not be generalizable to the whole population. As a result, the magnitude of socioeconomic effects on PRS performance may differ in other populations and healthcare systems.

Future work could incorporate breast cancer molecular subtype and finer-resolution socioeconomic measures, which we did not consider here due to their unavailability in the All of Us Research Program. Breast cancer subtype is particularly relevant for risk prediction because triple-negative breast cancer disproportionately affects Black American women and contributes substantially to disease burden and mortality (Prat et al., 2015). In addition, neighborhood socioeconomic status is positively associated with the incidence of aggressive breast cancer subtypes, including triple-negative breast cancer (Aoki et al., 2021; Qin et al., 2021). All of Us v8 measures deprivation index at a three-digit ZIP code resolution, far broader than neighborhood level data. Accounting for these factors could give insight into subtype-or neighborhood-specific genetic and environmental effects.

Despite the promise of PRS for cancer risk stratification, low predictive accuracy and equity remain barriers to clinical implementation. Existing genomic resources disproportionately represent individuals of European ancestry and higher socioeconomic status, and deployment of current polygenic scores without careful evaluation risks exacerbating existing health disparities. Our findings indicate that socioeconomic status influences breast cancer PRS transferability within and across racial groups and contributes independently to breast cancer risk prediction. Future studies should therefore consider socioeconomic context as part of the equitable evaluation and deployment of polygenic risk scores.

## Data Availability

The individual-level genome-wide, survey, and phenotypic data used in this study are publicly accessible to registered researchers through the All of Us Researcher Workbench (https://workbench.researchallofus.org/) following Data and Research Center (DRC) access procedures. Genomic data (v8 release) and survey responses, including the Basics survey, are part of the All of Us controlled tier dataset. Any additional code used in this study will be made available upon request to registered All of Us researchers via the Research Workbench.

## Acknowledgements

We gratefully acknowledge the All of Us participants for their contributions, without whom this research would not have been possible. Thank you to our colleagues in the Musharoff lab for providing constructive discussion and feedback, especially Jaclyn Liquori. This research was supported by NIH U54CA267738 (SAM).

## Supplementary Figures

**Supplementary Figure 1.**
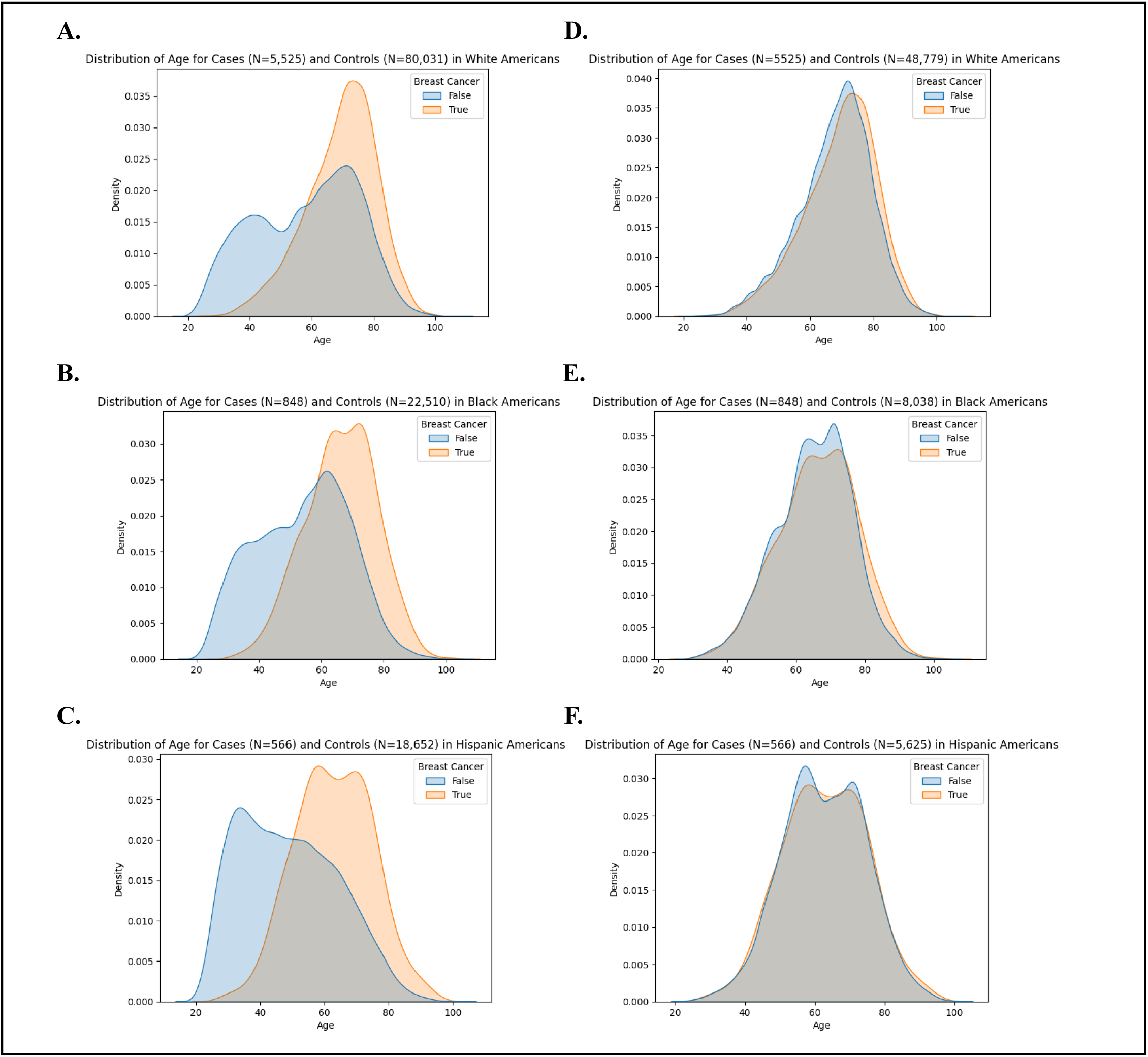
Selecting ancestry-specific age-matched controls. For each group: (A) White, (B) Black, (C) Hispanic, age bins of length 5 were constructed, ranging from 20 to 110 (case ages range: 23-106 years). For each case within a specific bin, 10 controls with matching self-identified race and ethnicity as the case were selected without replacement. Results of this age matching are also shown (D) White, (E) Black, (F) Hispanic.

**Supplementary Figure 2.**
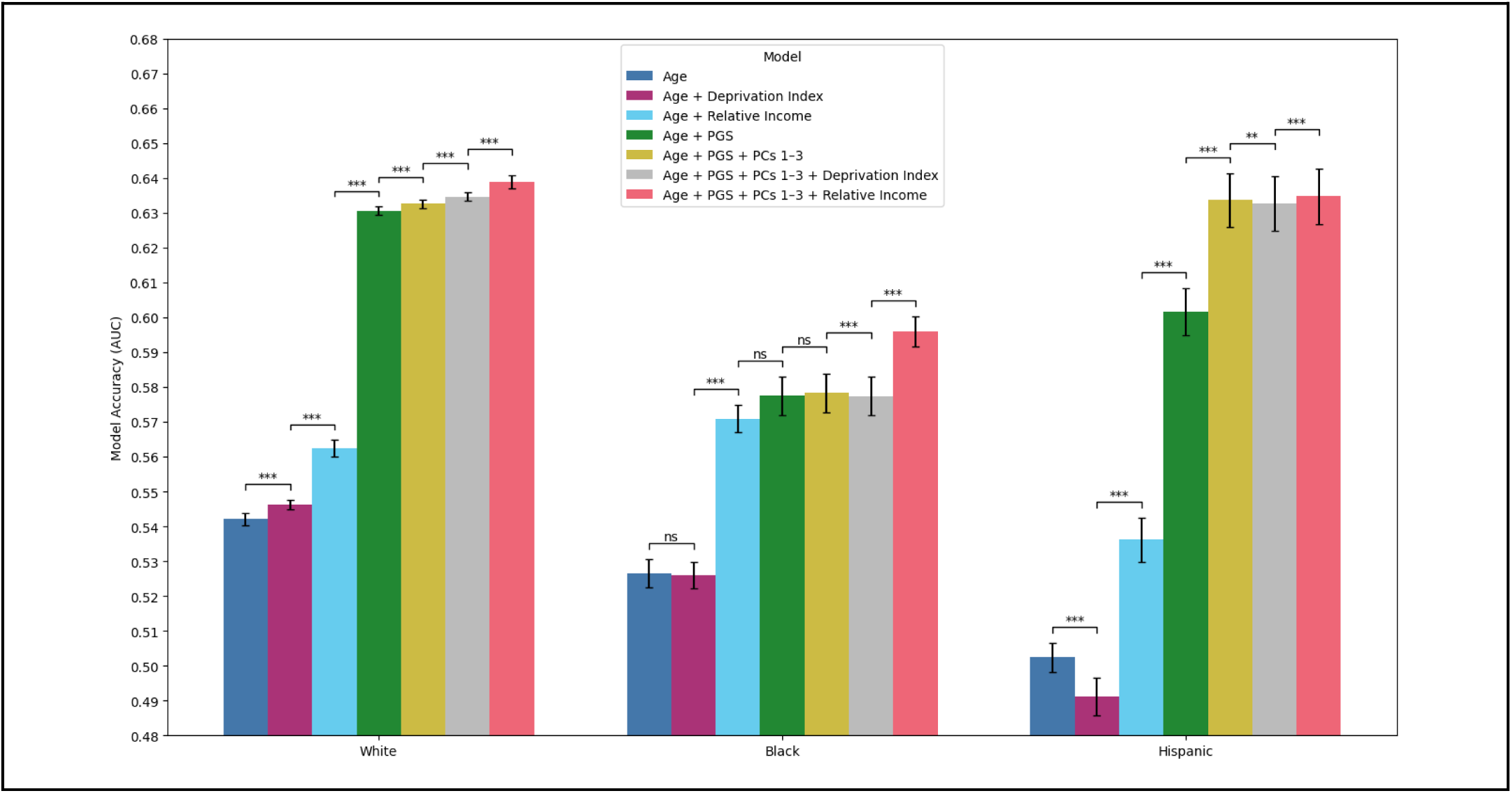
The addition of a 3-digit zip code deprivation index has minimal impact on breast cancer prediction models. Bars represent 5-fold cross-validated AUCs with a 1:5 case to control ratio across 10 independent repetitions for White Americans (left), Black Americans (middle), and Hispanic Americans (right), sampled from an age-matched dataset [Supplementary Figure 1]. Models are sequentially additive: Age; Age + Deprivation Index; Age + Relative Income; Age + PRS; Age + PRS + Genetic PCs 1-3; Age + PRS + PCs 1-3 + Deprivation Index; and Age + PRS + PCs 1-3 + Relative Income. The PRS uses previously published weights without recalibration. Error bars denote 95% confidence intervals around the mean AUC, and 1-3 significance stars representing cutoffs of p < 0.05, p < 0.01, and p < 0.001 via paired t-tests comparing adjacent models within each racial group.

**Supplementary Figure 3.**
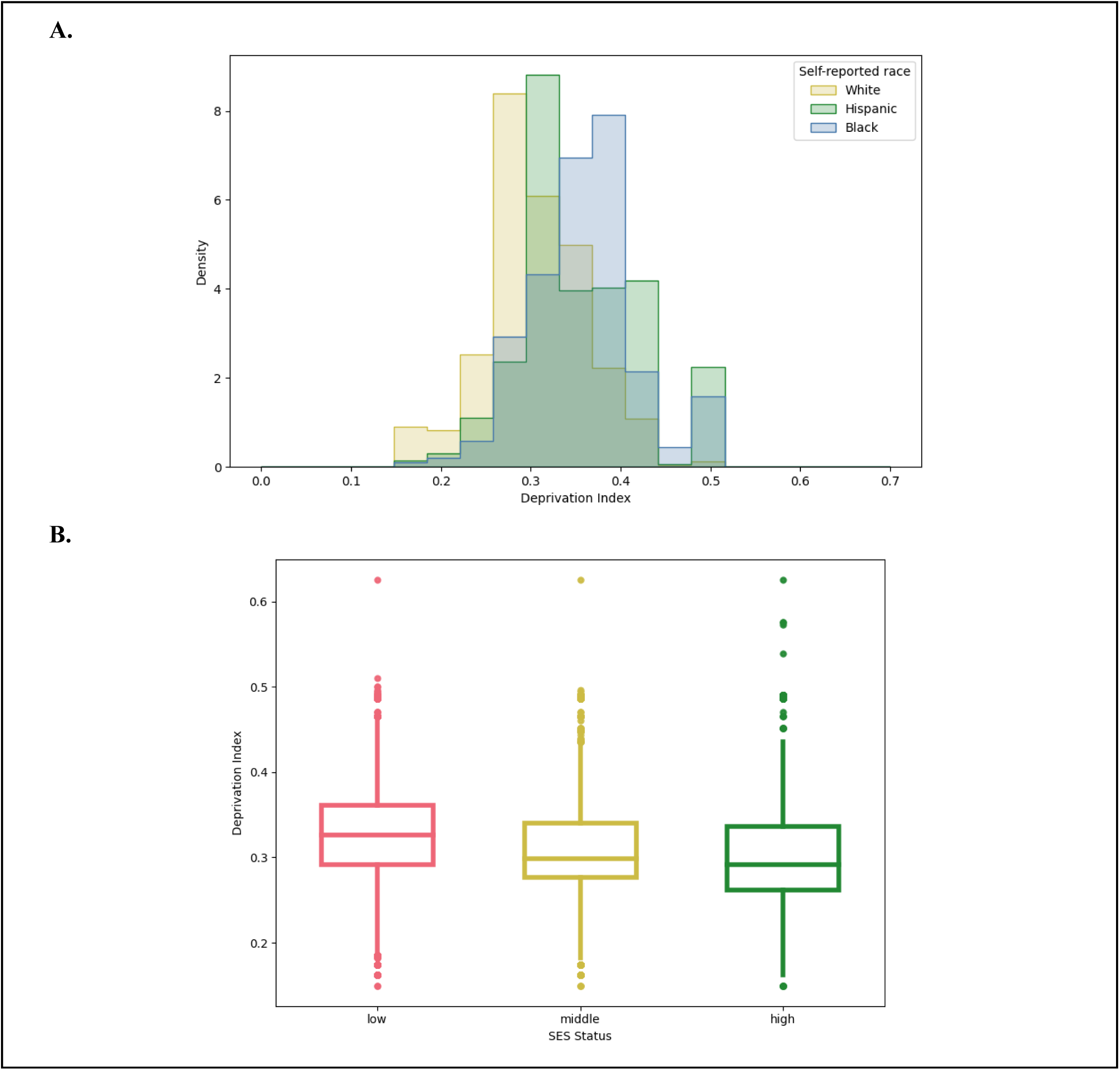
(A) Three-digit zip code neighborhood level deprivation index stratified by self-reported race is lower in White cohort compared to the other two cohorts. 99,325 White, 32,808 Hispanic, and 31,582 Black/AA participants. (B) Distribution of deprivation index stratified by socioeconomic status (SES). We summarized deprivation index across SES categories stratified by household income relative to the federal poverty line: low <300% (left panel); middle as 300%-600% (middle panel); and high as >600% (right panel).

**Supplementary Figure 4.**
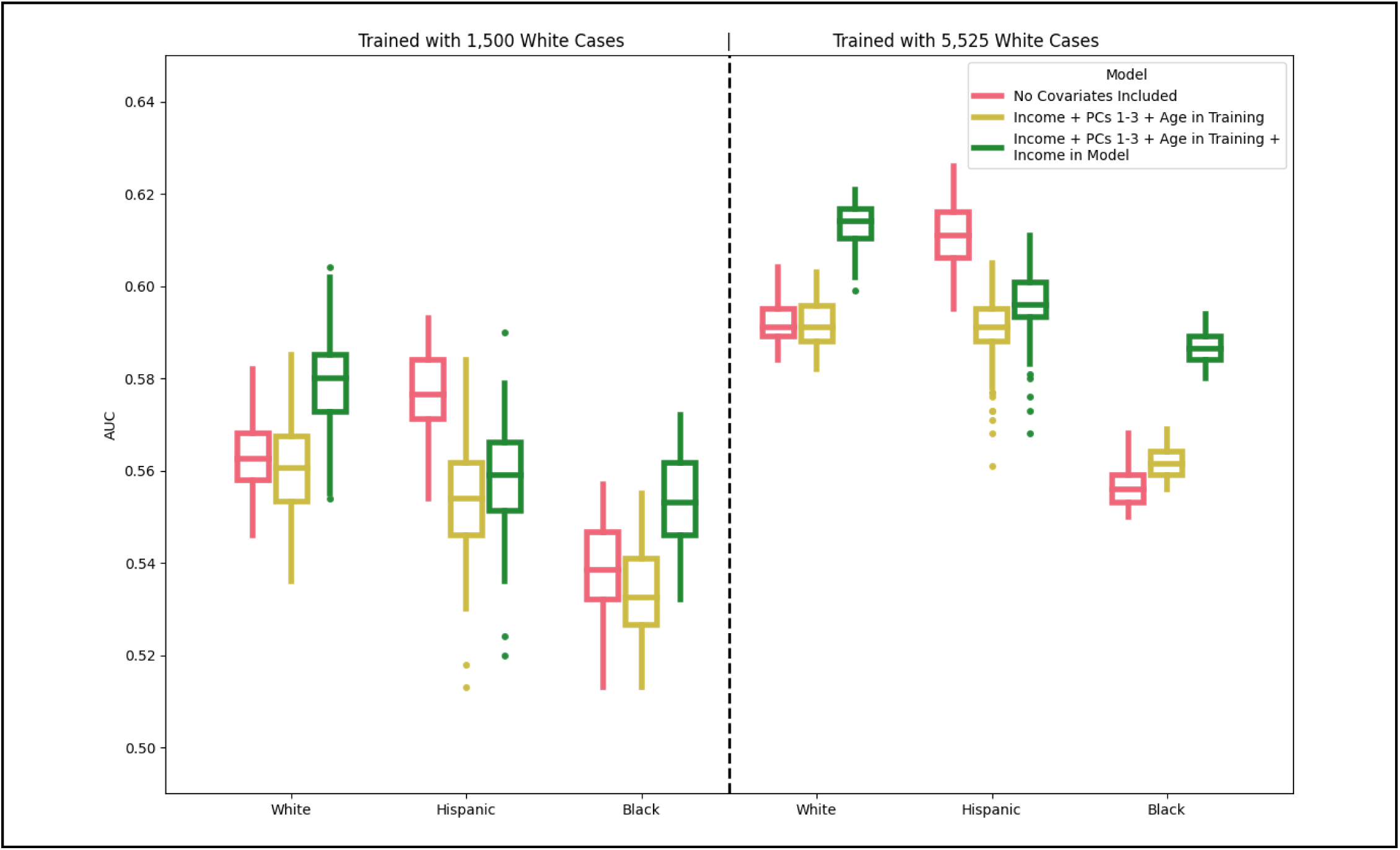
Adjustment for age, relative income, and genetic ancestry during PRS recalibration affects breast cancer PRS performance across racial groups. PRSs were recalibrated in White Americans and evaluated in White, Hispanic, and Black American validation cohorts. Five-fold cross-validation was performed using an 80:20 training-validation split with 10 bootstrap repeats, yielding 50 evaluations per group. PRSs were trained in White Americans using either 1,500 cases and 15,000 controls (left) or 5,525 cases and 80,031 controls (right), then evaluated in White, Hispanic (566 cases, 18,652 controls), and Black (848 cases, 22,510 controls) validation cohorts. Models included PRS only (red), PRS recalibrated with age, relative income, and genetic PCs 1–3 included as covariates during variant effect-size estimation (yellow), and PRS recalibrated with age, relative income, and genetic PCs 1–3 during variant effect-size estimation plus relative income included as a covariate in the final prediction model (green).

**Supplementary Figure 5.**
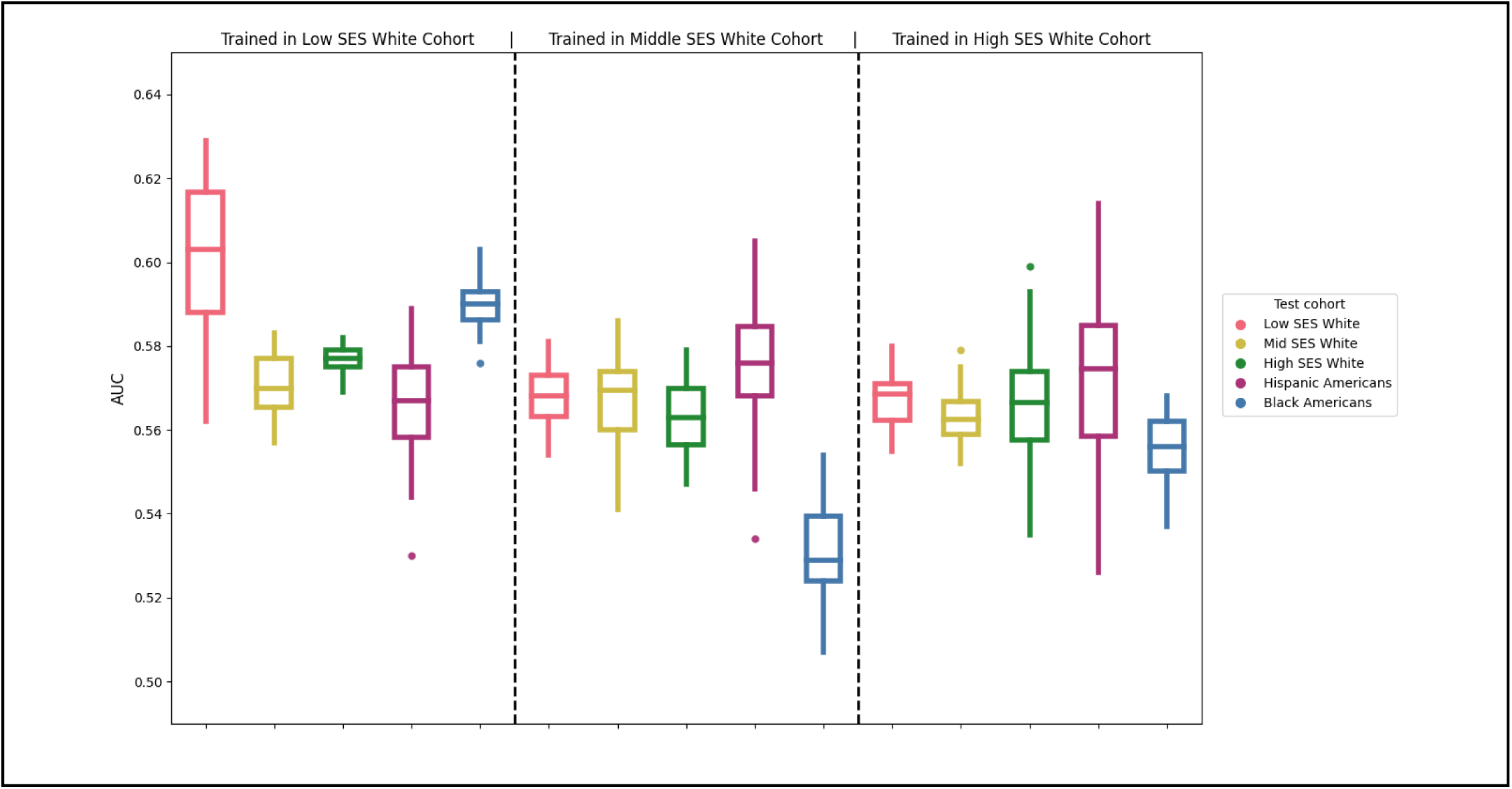
The transferability of White-derived PRS varies by socioeconomic status, with additional covariates included during variant effect size estimation. White individuals were stratified by household income relative to the federal poverty line into low (<300%; left), middle (300%-600%; middle), and high (>600%; right) SES groups. For each stratum, PRSs were trained using 5-fold cross-validation (1:5 case to control ratio) with age, relative income, and genetic PCs1-3 included as a covariate during model fitting to estimate variant effect sizes. The resulting SES-stratified PRSs were evaluated in held-out White cohorts stratified by SES and in unstratified Hispanic (566 cases, 18,652 controls) and Black (848 cases, 22,510 controls) cohorts, with relative income included as a covariate in the prediction model. The white SES-stratified training groups consisted of 1,500 cases and 15,000 controls.

**Supplementary Figure 6.**
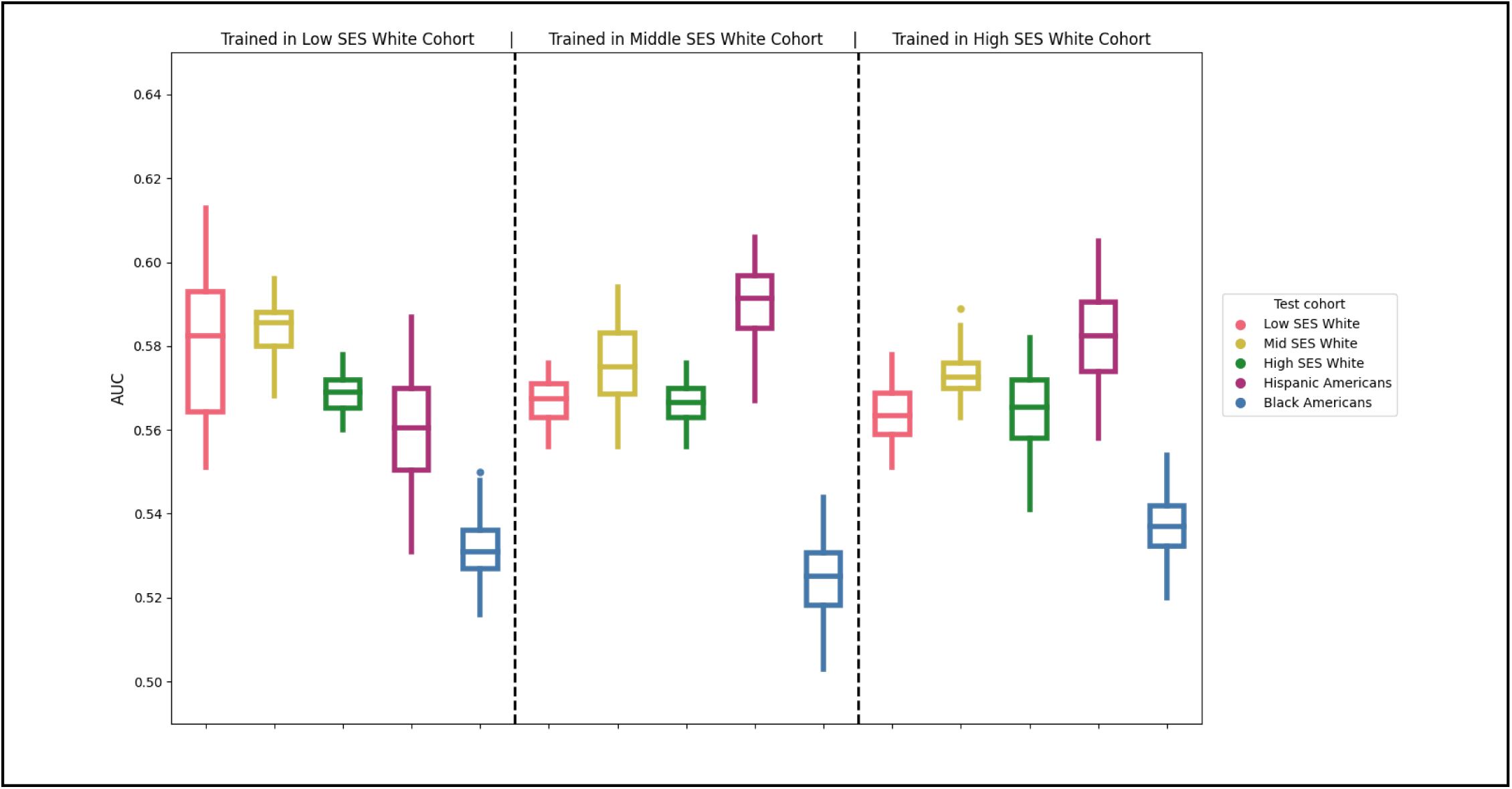
The transferability of White-derived PRS varies by socioeconomic status, without income as a covariate in the final model. White individuals were stratified by household income relative to the federal poverty line into low (<300%; left), middle (300%-600%; middle), and high (>600%; right) SES groups. For each stratum, PRSs were trained using 5-fold cross-validation (1:5 case to control ratio) without covariates in model fitting to estimate variant effect sizes. The resulting SES-stratified PRSs were evaluated in held-out White cohorts stratified by SES and in unstratified Hispanic (566 cases, 18,652 controls) and Black (848 cases, 22,510 controls) cohorts. The white SES-stratified training groups consisted of 1,500 cases and 15,000 controls.

**Supplementary Figure 7.**
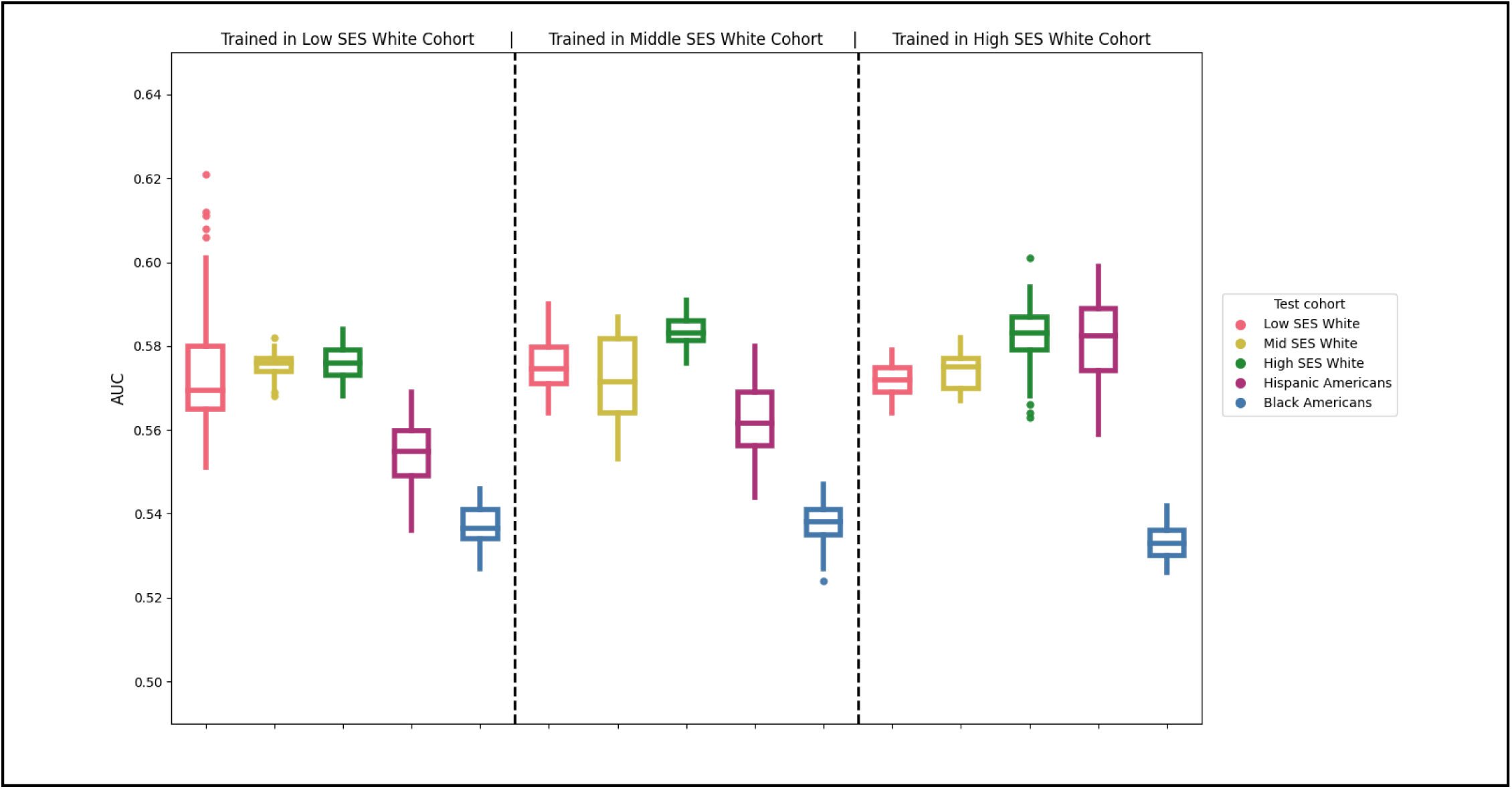
Three-digit zip code deprivation index stratification. Models were trained in White individuals stratified by 3-digit zip code deprivation index: low < 0.28 (left panel); middle as 0.28-0.33 (middle panel); and high as >0.33 (right panel). Each training group (low, middle, and high) consisted of 2,000 cases, 20,000 controls. SES-stratified PRSs were trained in White individuals using 5-fold cross-validation bootstrapped 10 times with a 1:5 case-to-control ratio to produce recalibrated weights. We then applied the resulting PRS across White Americans (low-, middle-, and high-SES), Hispanic Americans (566 cases, 18,652 controls), and Black Americans (848 cases, 22,510 controls).

